# Whole genome sequencing and transmission analysis of *Vibrio cholerae* isolates from Eastern and Southern Africa: a genomic epidemiology study

**DOI:** 10.1101/2024.03.28.24302717

**Authors:** Shaoming Xiao, Ahmed Abade, Waqo Boru, Watipaso Kasambara, John Mwaba, Francis Ongole, Mariam Mmanywa, Nídia Sequeira Trovão, Roma Chilengi, Geoffrey Kwenda, Christopher Garimoi Orach, Innocent Chibwe, Godfrey Bwire, O. Colin Stine, Aaron M. Milstone, Justin Lessler, Andrew S. Azman, Wensheng Luo, Kelsey Murt, David A. Sack, Amanda K. Debes, Shirlee Wohl

## Abstract

**Background:** Despite ongoing containment and vaccination efforts, cholera remains prevalent in many countries in sub-Saharan Africa. Part of the difficulty in containing cholera comes from our lack of understanding of how it circulates throughout the region, so this study uses genomic epidemiology to identify disease transmission patterns in Southern and Eastern Africa.

**Methods:** To better characterize regional transmission, we performed whole genome sequencing on 142 *Vibrio cholerae* samples of different sample types, each collected between 2007-2019 from five different countries in Southern and Eastern Africa. We obtained 114 high quality *V. cholerae* genomes that we combined with 1385 previously published genomes to conduct phylogenetic and other analyses used to better understand cholera transmission and circulation in Southeastern Africa.

**Findings:** We showed that *V. cholerae* sequencing can be successful from a variety of sample types and filled in spatial and temporal gaps in our understanding of circulating lineages, including providing some of the first sequences from the 2018-2019 outbreaks in Uganda, Kenya, Tanzania, and Malawi. Our results present a complex picture of cholera transmission in the region, with multiple lineages found to be co-circulating within several countries.

**Interpretations:** Our findings suggest that previously identified sporadic cases may be from larger, undersampled outbreaks, highlighting the need for careful examination of sampling biases and underscoring the need for continued and expanded cholera surveillance across the African continent.

**Funding:** Funding for this project was provided by the National Institutes of Health and the Bill and Melinda Gates Foundation.

**RESEARCH IN CONTEXT:** *Evidence before this study:* A comprehensive meta-analysis of *Vibrio cholerae* O1 published in 2017 identified 12 introductions of *V. cholerae* from Asia into Africa, designated as T1-T12 (or AFR1-AFR12). More recently, a study published in 2019 surveyed *V. cholerae* O1 from the 2016-2017 outbreak in Yemen and identified a 13th introduction, and studies from 2022 and 2023 identified the T14 and T15 sublineages, respectively. Existing *V. cholerae* genomic data includes sequences from the two publications referenced above, as well as 15 other publications referenced in that manuscript, which provide a representative set of *V. cholerae* O1 in the African continent and globally over time. Using PubMed searches for terms including (“Vibrio cholerae”) and (“Africa”) and (“Sequencing or Genomics”), we identified an additional four studies with publicly available genomic data from Africa, published after the genomic studies mentioned above and before July 2021. Although the initial cholera genomics efforts captured the continent-level landscape of *V. cholerae* diversity and more recent studies describe the transmission of *V. cholerae* within specific countries, few studies use genomic data to explore regional, multi-country transmission patterns on the African continent.

*Added value of this study:* This study generated 114 *V. cholerae* O1 genomes from samples collected in Kenya, Tanzania, Uganda, Malawi and Zambia from 2007-2019, allowing us to take a regional look at cholera transmission in Southern and Eastern Africa. Joint analysis of genomes from several different countries allowed us to better understand patterns of spread, including the potential emergence of new sublineages, and helped fill gaps in our understanding of how cholera moves on the African continent. Additionally, whereas most *V. cholerae* genomics studies rely on bacterial isolates for whole genome sequencing, we generated sequences from multiple sample types, indicating that commonly used low-cost sample preservation methods may be useful for genomic studies.

*Implications of all the available evidence:* Data generated from this study suggest a more complex picture of *V. cholerae* transmission in Southern and Eastern Africa than previously thought, and the need to carefully consider sampling limitations in our interpretation of *V. cholerae* genomic data. Additionally, our results highlight the importance of a coordinated, regional approach to cholera surveillance.

## INTRODUCTION

Cholera is an acute watery diarrheal disease that is estimated to cause approximately 95,000 deaths annually in endemic countries ^1^. Most of these deaths occur during large outbreaks that are associated with specific serotypes of the *Vibrio cholerae* bacterium—O1 and O139. The ongoing seventh pandemic of cholera, caused by *V. cholerae* serogroup O1 biotype El Tor (and often referred to as “7PET”, for 7th Pandemic El Tor), was first detected in Indonesia in 1961 and has since spread through Asia to Africa, Europe, and Latin America ^2^. Africa, especially sub-Saharan Africa, has experienced a higher cholera burden during the seventh pandemic than other regions of the world ^1,3^, with numerous large outbreaks reported since 1961, including several during the last decade ^4^. Although a few studies have explored the transmission patterns of *V. cholerae* in Africa, there is still a limited understanding of how cholera circulates ^5^.

Unlike endemic regions in Southeast Asia, cholera in Africa is often seasonal and periodic, with many countries experiencing months or years of no cholera cases between successive outbreaks. Although risk factors such as population density, seasonality and urbanization may explain some of variability in observed transmission patterns ^2,6–8^, within-country variation in some of these factors (e.g., seasonality) ^9^ highlights the limitations of these risk factors in fully capturing cholera dynamics. For example, most cholera outbreaks in Malawi have historically occurred during the rainy season, but the 2022 outbreak (which has caused approximately 60,000 cases as of September 2023 ^10^) started in the dry season ^11^. It is clear that these factors alone cannot explain the repeated reemergence of cholera in regions without ongoing cases, and that fully elucidating cholera dynamics on the continent will require a better understanding of *V. cholerae* movement within and between countries. Importantly, filling gaps in our understanding of the transmission patterns of *V. cholerae* has the potential to improve how we prepare and respond to outbreaks, in part by informing where to address containment, vaccination, and other outreach efforts—some of which could be most effective when implemented across country borders.

Genomic analysis provides some insight into the movement of pathogen lineages and has been used already to track *V. cholerae* O1 transmission in Africa. Early genomic studies showed that 7PET *V. cholerae* O1 was introduced from Asia to Africa at least 12 times since 1970 ^12^. These introduction events occurred mainly in West and Southeast Africa, each defined by a specific sublineage (AFR1-AFR12) ^13^. At least three additional lineages (AFR13, AFR14, and AFR15) ^12–15^ have been identified more recently, all following the same Asia-to-Africa introduction pattern. More recent lineages have generally appeared to replace older lineages upon introduction ^12,14^, though there is still limited information about how these lineages interact and circulate in sub-Saharan Africa once present.

A number of more recent studies have used genomics to characterize *V. cholerae* in Southeast Africa ^16–20^, a region of particularly high cholera burden ^1,3^. However, the difficulty of sequencing cholera in low-resource settings ^21^ means there are still significant gaps in our understanding of which lineages were circulating at different types over the last few decades. Furthermore, there have been limited opportunities to explore potential transmission across country borders ^22^. In this study, we aimed to fill some of the gaps in our understanding of 7PET *V. cholerae* O1 circulation in Southeast Africa using isolates collected from 2007– 2018 in Uganda, Kenya, Tanzania, Malawi, and Zambia. In addition, we explored the potential of performing whole genome sequencing from low-cost sample preservation methods (e.g., stool or isolates on filter paper), which facilitates specimen collection in resource-constrained areas where isolates would otherwise not be captured ^16,23^.

## METHODS

### Study design and bacterial isolates

#### Sample collection

Samples in all countries were collected as part of outbreak investigations from 2007-2019 (Kenya); 2016-2018 (Tanzania); 2017-2018 (Uganda); 2016-2018 (Zambia and Malawi) by the relevant National Public Health Laboratories. In Malawi, sample collection was restricted to patients with a known antibiotic consumption history. In order to confirm cholera from suspected cases in each country, specimens were first tested by cholera rapid diagnostic tests (RDTs) (Crystal VC, Arkray, Healthcare Pvt Ltd., Surat, India) or by Cholkit Ag O1 RDT (Incepta Pharmaceuticals Ltd, Bangladesh). Specimens that produced RDT positive tests were preserved in Cary Blair transport media sent from peripheral health facilities to local and/or regional laboratories and, with the exception of Malawi, spotted directly onto Whatman 903 Protein Saver Card (GE Healthcare Ltd., Forest Farm, Cardiff, UK).

For microbiological confirmation, specimens were streaked directly onto Thiosulfate Citrate Bile Salt sucrose (TCBS) agar and incubated as described in Supplementary Data 1. Cholera-like colonies were then tested for evidence of *V. cholerae* and positives were preserved in agar or on filter paper, as described in Supplementary Data 1.

#### Genomic study design

From the cholera-confirmed specimens and isolates available for sequencing, we selected samples that maximized the number of distinct geographic districts, with the goal of obtaining the most comprehensive picture of cholera transmission possible. In Kenya, for example, we sequenced samples from 16 different regions. We also selected samples to focus on more recent outbreaks for which no or limited sequencing data were available, with the goal of filling in gaps in current studies. We continued sequencing until we were no longer finding new lineages or adding new districts or years to our dataset.

#### Ethics statement

Samples from Kenya were collected as part of studies approved by the relevant Institutional Review Boards or Ethics Committees at Johns Hopkins Bloomberg School of Public Health (JHSPH; IRB numbers: 00009067, 00008982) and locally (approval number: ESRC P552/2018). Samples in Zambia, Tanzania, and Uganda were collected under JHSPH IRB 00008193, and IRB00008221, respectively, with local approval. Samples analyzed from Malawi were collected under routine public health surveillance. Sequencing and secondary analysis of these samples were determined by JHSPH to be non-human subjects research and exempt from further review.

### Procedures

#### DNA extraction and quantification

Specimens from all countries were sent to Baltimore, USA for extraction and molecular analysis. Filter paper specimens (Kenya, Zambia, Uganda, Tanzania) were extracted by boiling and glycerol stock isolates from Malawi were extracted using the QIAamp DNA Mini Kit (Qiagen), as described in Supplementary Data 1. All extracted nucleic acid was confirmed using conventional PCR and quantified on the Qubit Fluorometer (Thermo Fisher) using the dsDNA High Sensitivity Kit according to instrument instructions.

#### Library construction and sequencing

Illumina library preparation and sequencing was performed in Baltimore, USA using the Nextera DNA Flex Library Prep kit (Illumina). Samples were pooled and sequenced on the Illumina NovaSeq platform with 2×150 bp paired-end reads. Oxford Nanopore library preparation and sequencing was also performed in Baltimore following the SQK-LSK109 library preparation kit (Oxford Nanopore Technologies), with minor modifications as described in Supplementary Data 1. Samples were run on the MinION run for 48 hours per run and the resulting data was basecalled using Guppy version 3.0.3 with model dna_r9.4.1_450bps_fast.cfg. Adapter removal and demultiplexing was performed as described in Supplementary Data 1.

#### Reference-based genome assembly

Illumina paired-end reads were aligned against *V. cholerae* O1 El Tor N16961 (accession: AE003852/AE003853) and assembled following the pipeline available here: https://github.com/HopkinsIDD/illumina-vc. For Oxford Nanopore data, reference-based genome assembly was performed as described in Ekeng et al. ^22^, full pipeline available here: https://github.com/HopkinsIDD/minion-vc.

### Statistical analyses

#### Maximum likelihood estimation

We combined the sequences generated in this study (n=114, see **Supplementary Data 1**) with 1,385 publicly available genomes (**Supplementary Data 2**) and masked recombinant sites as described in Weill et al. ^12^. We then used IQ-TREE version 1.6.12 to construct a maximum likelihood phylogeny from the full masked dataset of 1,499 genomes. Further details are available in Supplementary Data 1.

#### Bayesian phylogenetic reconstruction

Before performing Bayesian phylogenetic analysis, we removed 11 temporal outliers from our background dataset using TempEst ^24^ (**Supplementary Figure 1**). These outliers are likely due to sequencing errors or inaccurate sample collection dates. We then used BEAST version 1.10.5 ^25^ to estimate temporal dynamics of wave 3 sequences (n=961, see **Supplementary Data 2**). We created a maximum clade credibility tree (**Supplementary Figure 2**) and calculated the time to the most recent common ancestor (tMRCA) for lineage AFR13, which we report in **Figure 3** (95% highest posterior density (HPD) interval).

## RESULTS

To better understand cholera transmission in Southeast Africa, we performed whole genome sequencing (WGS) of *V. cholerae* isolates from five neighboring countries in this region: Uganda, Kenya, Tanzania, Malawi, and Zambia (**Fig 1A**). In total, we generated 114 high-quality genomes (4, 67, 22, 20, and 1 from Uganda, Kenya, Tanzania, Malawi and Zambia, respectively) from 142 sequenced isolates collected between 2007-2019 (**Supplementary Data 3**). These data fill in a number of geographic and temporal gaps in available *V. cholerae* genomic data and provide some of the first sequences from the 2018-2019 outbreaks in Uganda, Kenya, Tanzania, and Malawi (**Fig 1B**).

**Figure 1.**
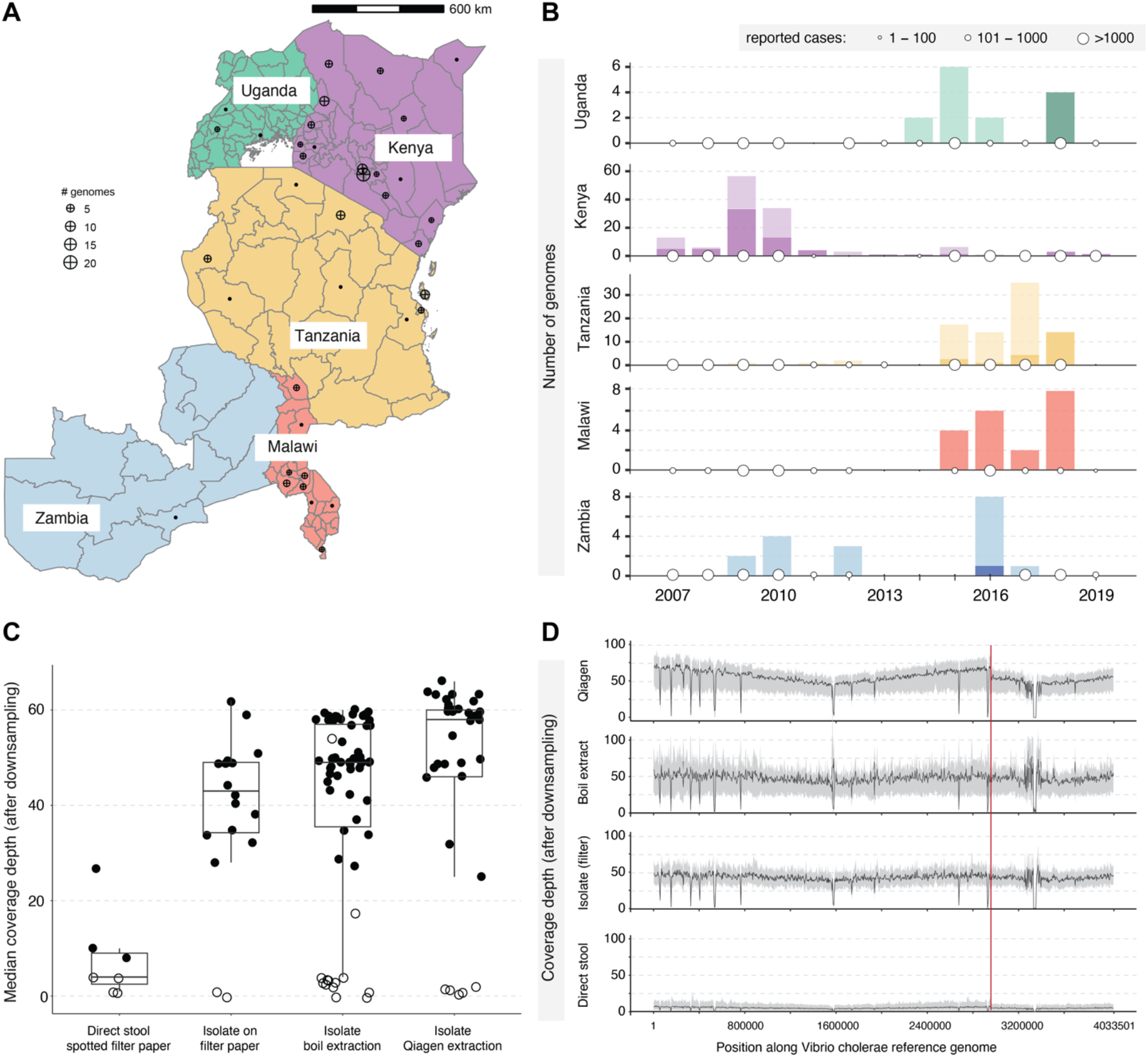
*Vibrio cholerae* sequences generated in this study. (**A**) Geographic distribution of genomes generated in this study. 9 samples from Kenya with no sub-country metadata are not shown. (**B**) High quality genomes generated in this study (dark bars) overlaid on previously published genomes (light bars), per country and per year (genomes with unknown collection year excluded) (see also **Supplementary Data 2, 3**). White dots on x-axis: total number of cholera cases reported to the World Health Organization in that year ^29^ (**Supplementary Data 4**). (**C**) Median coverage depth, per sample type and extraction method (all samples were boil extracted except those specifically marked as Qiagen extractions), across the *V. cholerae* genome after downsampling reads to 1 million/sample. Only samples sequenced on the Illumina platform are included. 2 samples of unknown sample type were not shown. Black dots: samples classified as high-quality genomes (based on non-downsampled reads; see **Methods**); white dots: low-quality genomes excluded from further analyses. Box plot outliers are those with median coverage >18.75 (direct stool spotted filter paper); >71.125 or <12.125 (isolate on filter paper); >89.25 or <3.25 (isolate boil extract); or >81 or <25 (isolate Qiagen extraction) (**Supplementary Data 5**). (**D**) Distribution of coverage depth at each position along the *V. cholerae* reference genome for each sample preparation after downsampling reads to 1 million/sample. Solid black line: median coverage across all samples within the sample type group; gray bands: 20-80 percentile range, averaged across a 4000-nt sliding window. (**Supplementary Data 6**) Red vertical line: boundary between chromosomes 1 and 2 of *V. cholerae* genome.

Other than sequencing costs, one of the barriers to generating *V. cholerae* whole genome sequences is heterogeneity in the sample types available; sequencing typically requires relatively large amounts of purified nucleic acid, and is therefore often not attempted when only crude specimens (as opposed to cultured isolates) are available. Previous studies have shown that *V. cholerae* sequencing from Alkaline Peptone enriched (APW) stool or isolates on filter paper can be successful ^23^, so we attempted to take this one step further, comparing the success of various specimen types preserved on filter paper across samples from several countries and using different nucleic acid extraction techniques. Specifically, we compared sequencing from bacterial cultures, bacterial isolates preserved on filter paper, and whole stool directly spotted onto filter paper. We experimented with both column-based extraction kits as well as boil extraction methods, which can be done at reduced cost, with less laboratory equipment, and (when isolates are preserved on filter paper) without temperature-controlled supply chains.

As expected, we found that, after normalizing the total number of reads per sample (see **Methods**), column-extracted isolates produced the highest quality *V. cholerae* genomes, while direct stool spotted filter paper samples had the lowest median coverage depth across the *V. cholerae* genome (**Fig 1C**). To minimize potential platform-related effects, these results include only samples sequenced on the Illumina sequencing platform (all samples except those collected in Malawi, see **Supplementary Data 3**). Overall, the sequences from all specimen preservation types produced high quality genomes that met our inclusion criteria (see **Methods**). Although the direct stool on filter paper had the lowest median coverage, sample size for this sample type was quite small. Three of seven samples of this type still produced high quality genomes according to our thresholds, without any modification to the sequencing method used.

To ensure that our conclusions about sequencing quality were not driven by highly uneven coverage, we also looked at read depth across the whole *V. cholerae* genome (**Fig 1D**), again focusing only on samples sequenced on the Illumina platform for ease of comparison. We observed relatively even coverage across the genome, with similar trends across all sample types.

Using high quality genomes from all sample types, we then built a maximum likelihood tree using the 114 high quality genomes generated in this study and 1,374 previously published genomes (**Supplementary Figure 1, Supplementary Data 2** for details and acknowledgement). Using this phylogeny, we determined that sequences generated in this study fell within the AFR10, AFR11 and AFR13 lineages (**Fig 2, Fig 3A**).

**Figure 2.**
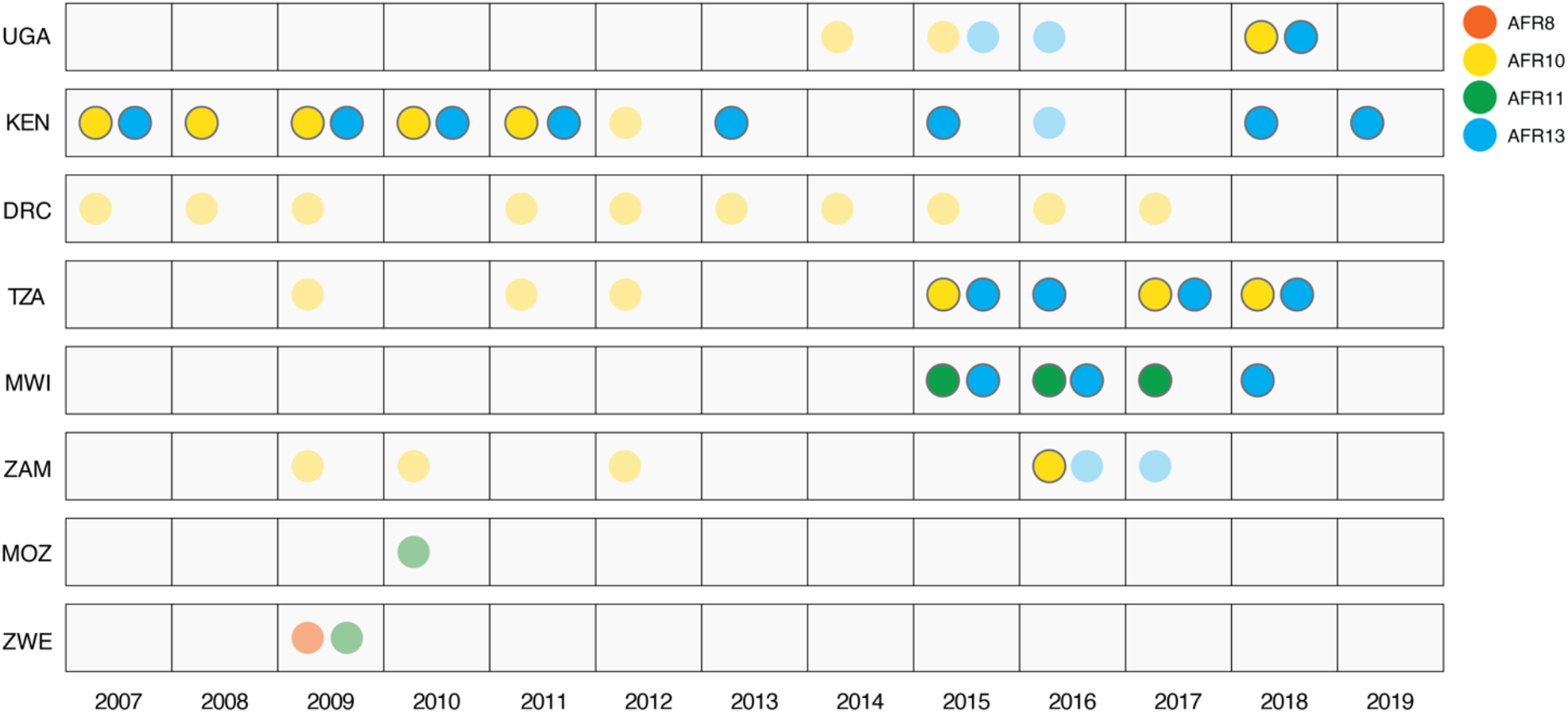
Distribution of lineages identified in samples from this study. Countries in Eastern and Southern Africa, ordered from North to South. Outlined circles: lineages found in genomes from this study. Faded circles (no outline): lineages found in previously published genomes. Abbreviations: UGA (Uganda), KEN (Kenya), DRC (Democratic Republic of the Congo), TZA (Tanzania), MWI (Malawi), ZAM (Zambia), MOZ (Mozambique), ZWE (Zimbabwe).

**Figure 3.**
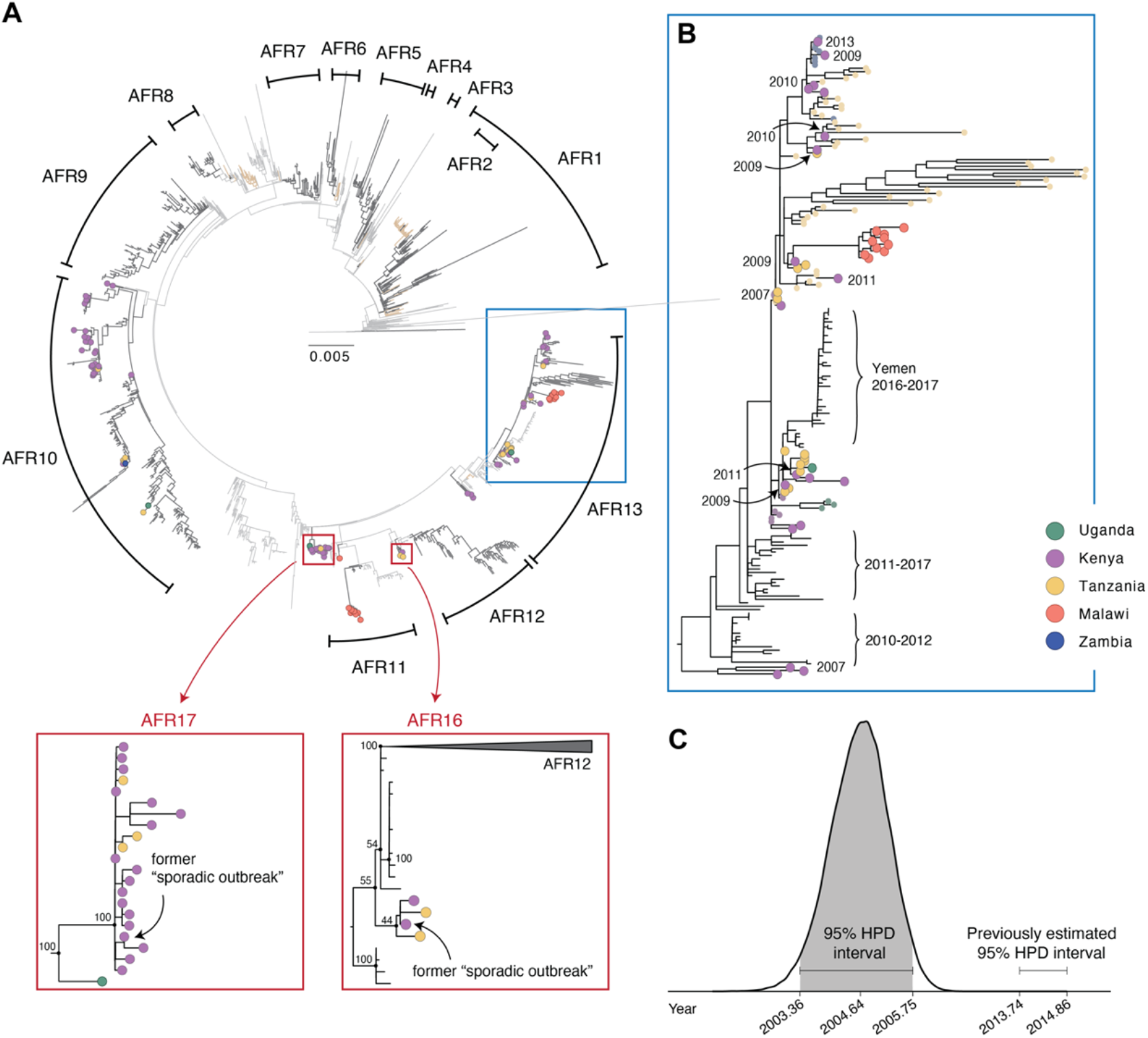
Phylogeny of *V. cholerae* O1 sequences. (**A**) Maximum likelihood tree of 1488 *V. cholerae* genomes (see also **Supplementary Data 7**) Colored tips: 114 genomes generated in this study, colored by country of sample collection. Branches are colored by continent: Asia (light gray), Africa (dark gray), all other areas of the world (light brown). Proposed AFR16 and AFR17 nomenclature shown in red (AFR14 and AFR15 were previously identified but in samples more recent than the study period used here, and therefore are not shown). (**B**) Zoom view of the blue box area shown in (A) showing the AFR13 lineage. All African sequences from prior to 2015 are labeled with the year of collection, and all tips without a year label are from samples collected in 2015-2018. Colored tips: genomes from African countries. Larger tips: genomes generated in this study; smaller tips: previously published genomes from African countries. (**C**) Posterior distribution of time of the most recent common ancestor (tMRCA) of the AFR13 lineage given the dataset presented in this manuscript, as compared to previous estimates ^12^ (see **Supplementary Data 8**).

Most of these sequences belong to lineages previously identified in the country of sample origin, though we also observed some unexpected results, suggestive of a more complex picture of cholera transmission than previously thought.

First, our data highlight that two strains previously considered to be “sporadic outbreaks” because they did not fall into one of the known AFR lineages may reflect undersampled, larger outbreaks. In both cases, these strains were previously isolated on the phylogenetic tree, suggesting they may have not spread widely within Africa once introduced ^12^. However, several newly generated genomes cluster closely with these two “sporadic” cases in our phylogeny (**Fig 3B**): 3 sequences from Kenya and Tanzania collected between 2009–2018 cluster closely with a former singleton from Kenya, and 21 genomes from Kenya, Tanzania and Uganda collected between 2009–2018 cluster closely with a separate former singleton. We have designated these sequences as AFR16 and AFR17, respectively.

Additionally, although the AFR13 sub-lineage has been found in East Africa in recent years (specifically Kenya, Tanzania, Uganda and Zimbabwe, as recently as 2019) ^14,16,17,19,26^, we found evidence that the introduction of this lineage into Africa may have been earlier than previously thought. The previously reported AFR13 sequences were from samples collected in Kenya, Tanzania and Uganda in 2015 ^14,17,19,27^, and these studies estimated that this lineage emerged between 2013-2014 ^14^. However, we generated 10 sequences from isolates collected in Kenya that fell within the AFR13 lineage but were collected between 2007-2011. These sequences are highly similar to others previously collected in the region and appear ancestral to sequences collected from the major outbreak in Yemen during 2016 to 2017 ^14^ (**Fig 3C**), thus suggesting the AFR13 lineage may have been circulating in Southeast Africa since at least 2007. Using a Bayesian phylogenetic analysis (**Supplementary Figure 2**), we confirmed that the estimated emergence of the AFR13 lineage is significantly earlier than previously estimates following the addition of the sequences generated in this study (2004.64, 95% highest posterior density (HPD) interval: 2003.36-2005.75) (**Fig 3D**).

We attempted to confirm these results by validating both the sequence data itself and the accompanying temporal metadata. All ten AFR13 sequences in question had high coverage depth and no evidence of contamination. Upon cross-checking sample collection dates, we also noted that the isolates were brought to the United States (where the nucleic acid was extracted and they were ultimately sequenced) prior to 2012 ^28^, so presence of AFR13 genomic data in these samples supported our conclusion, regardless of specific collection dates. Although it will be important to see if future genomic studies support this finding, we are including these sequences in our results because the release of these data could potentially be useful to the understanding of ongoing outbreaks.

Other sequences generated in this study (from multiple different countries) show the presence of co-circulating *V. cholerae* lineages in multiple Southeast African countries, suggesting complex relationships between the spread of different *V. cholerae* strains. Specifically, we observed multiple *V. cholerae* lineages in the same country within a single year in all five countries (**Fig 2, Supplementary Data 2**). For example, we observed multiple lineages circulating within Malawi in 2015 (**Fig 4A**) and 2016 (**Fig 4B**). To our knowledge, our data presents the first examples of multiple lineages persisting within a country in Eastern Africa in consecutive seasons, a phenomenon we also observed in Tanzania in 2017 and 2018 and Kenya in 2009-2011. That said, there are many countries and regions that reported cholera cases but have no genomic data available, so co-circulation may be more common than shown here.

**Figure 4.**
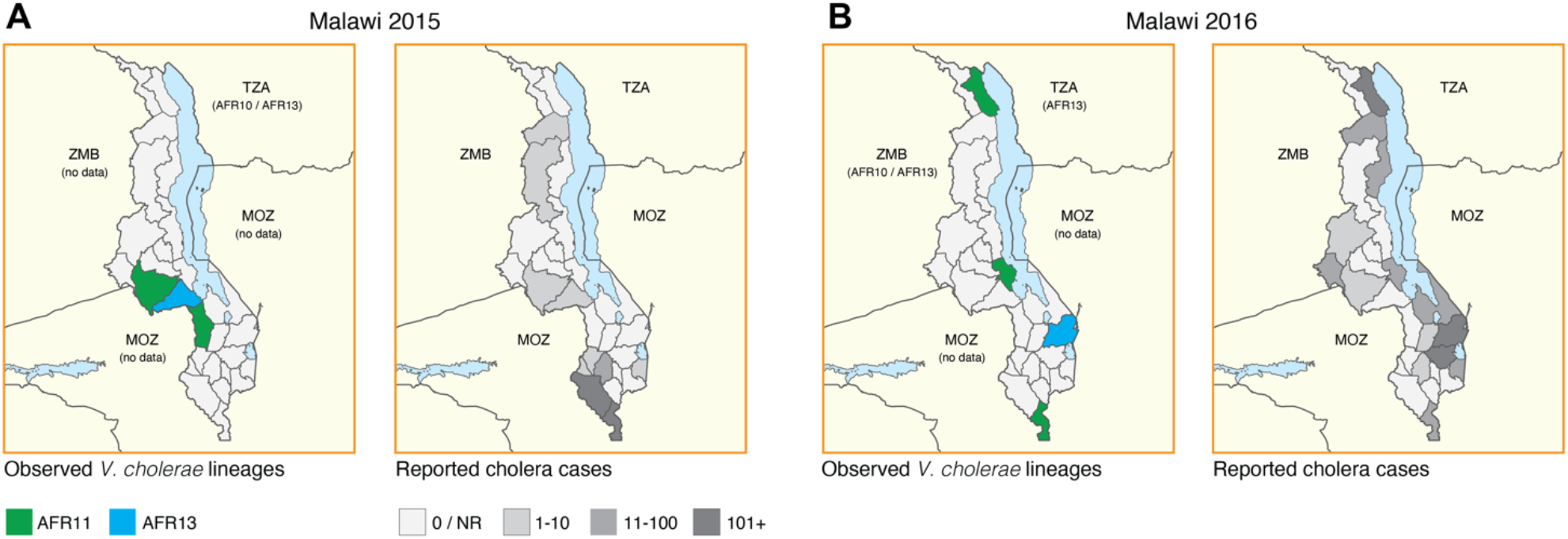
Co-circulation of multiple lineages at country or sub-country level. Examples of co-circulating lineages and movement of *V. cholerae* lineages (left) and reported cholera cases (right) in (**A**) 2015 and (**B**) 2016. Cholera cases from Kenya are as reported in Mutonga *et al*.^*30*^ and reported cholera cases in Malawi are as reported to WHO^29^ (**Supplementary Data 4**).

## DISCUSSION

In this study, we demonstrate that whole genome sequences of *V. cholerae* can be generated from stool and other non-culture sample types, and we use the resulting data to present an updated picture of cholera transmission. Specifically, we found evidence for undersampled lineages (AFR16 and AFR17), extensive co-circulation of multiple *V. cholerae* lineages within countries, and evidence supporting a revised understanding of when the AFR13 lineage was introduced to the African continent. Taken together, these findings suggest that improved genomic surveillance of cholera may yet uncover new aspects of cholera transmission in Southeast Africa. They also highlight that insufficient or biased sampling can lead to an inaccurate (as in the case of sequences now associated with AFR16 and AFR17) or incomplete picture of transmission—a potential issue for genomic studies of any pathogen that should be addressed with careful assessment of the potential effects of biased or under-sampling.

Given the frequent movement of *V. cholerae* from Southeast Asia to Southeast Africa, addressing these sampling biases will require that genomic surveillance of cholera improves simultaneously on both continents. For example, the addition of 10 *V. cholerae* sequences from African countries in this manuscript moved the estimated time to the most recent common ancestor of the AFR13 lineage to 2004, approximately 10 years earlier than previous estimates. However, the emergence of the lineage—an understanding of which could provide clues into *V. cholerae* transmission and evolution patterns within and between outbreaks—could have happened in either Africa or Asia, and additional sampling may further refine these date estimates. Improved sampling of both regions would also allow for the application of more complex phylodynamic methods for ascertaining the date and location of lineage emergence and an improved understanding of which lineages lead to local spread (as in the case of AFR16 and ARF17), though this data would be most useful with higher resolution epidemiological information, such as specific case location.

We also need to address some of the challenges associated with generating these sequences. Decreasing reliance on culture prior to sequencing will lower the burden to real-time sequencing (e.g., by enabling use of low-cost preservation methods such as filter paper), and seems increasingly possible even with current sequencing techniques, as shown by our successful sequencing from multiple sample types. Our results strengthen prior evidence that WGS can be effective from isolates stored on filter paper and serve as a proof of concept that WGS is effective from stool samples spotted directly on filter paper, though further optimization will be required to ensure these methods are as cost effective and accessible as possible. As whole genome sequencing becomes more widespread, it will also be important to enable rigorous documentation and high sample quality. We have done our best here to confirm our results with limited access to the original cultures, and we have documented these challenges as a caution to future researchers.

We also note that additional research is necessary on the relative merits of Illumina and Oxford Nanopore sequencing, both of which are used here. While Oxford Nanopore data produces results consistent with prior data on the lineage level ^22^, we note that sequences generated on this platform tend to cluster tightly and on longer branches than observed in Illumina-generated sequences. For this reason, we have refrained from drawing conclusions about the specific mutations or sublineage dynamics observed in these sequences (all isolates from Malawi were sequenced on the Oxford Nanopore platform). The long read data produced by Oxford Nanopore may be especially useful for understanding complex variation often observed in bacterial genomes, so additional research is needed to understand the source of these longer branch lengths.

Taken all together, our findings suggest a complex picture of *V. cholerae* transmission in Southeast Africa, and frequent co-circulation of multiple combinations of lineages. These findings also emphasize the importance of a regional approach to cholera surveillance and ^22^, as outbreaks in neighboring countries are connected both temporally (e.g., spikes in cases occur around the same time; **Fig 1D**) and molecularly (e.g., sequences from *V. cholerae* in multiple countries are highly related). Our hope is that this information can be useful in aiding cholera containment and mitigation, which may require cooperation across country borders. Ensuring these data continue to have public health impact will also require increased sampling (ideally as part of routine, in-country surveillance efforts), which should be supported by further investigation into optimal sample size and selection, especially in resource-limited settings. Finally, our data highlight the importance of pairing genomic data with epidemiological information and coordination with local experts who can interpret both routine and surprising results and ensure they are effectively shared and communicated with stakeholders across the continent.

## Data Availability

Raw data for all sequenced isolates and specimens is available under NCBI BioProject accession: PRJNA616030. R code used to make figures is available here: https://github.com/HopkinsIDD/CholeraGenomics-AfricaSE. Illumina bioinformatics pipelines are available here: https://github.com/HopkinsIDD/illumina-vc. Oxford Nanopore bioinformatics pipelines are available here: https://github.com/HopkinsIDD/minion-vc.

https://github.com/HopkinsIDD/illumina-vc

https://github.com/HopkinsIDD/minion-vc

## ACKNOWLEDGEMENTS

We thank David Mohr for laboratory assistance and the original authors of the sequences used in our phylogenetic analysis (references provided in **Supplementary Data 2**). We also thank Elizabeth C. Lee for her help collating cholera case data. Funding for this project was provided by the National Institutes of Health under award number R01HS028634 (A.M.M), K24AI141580 (A.M.M.), and R0AI123422 (A.K.D); Bill and Melinda Gates Foundation INV-047156 (D.S., A.K.D., S.W.); and OPP1195157 (J.L. and S.W.). The opinions expressed in this article are those of the authors and do not reflect the view of the National Institutes of Health, the Department of Health and Human Services, or the United States government.

## CONTRIBUTORS

Conceptualisation: A.K.D, S.W.; Data curation: S.X., N.S.T., K.M., S.W.; Formal analysis: S.X., N.S.T., S.W.; Funding acquisition: N.S.T., A.M.M., J.L., A.S.A., D.A.S., A.K.D., S.W.; Investigation: A.A., W.B., W.K., J.M., F.O., M.M., R.C., G.K., C.G.O., I.C., G.B., A.K.D.; Methodology: K.M., A.K.D., S.W.; Project administration: A.K.D., S.W.; Resources: O.C.S., J.L., A.S.A., D.A.S., A.K.D.; Software: J.L., S.W.; Supervision: A.K.D., S.W.; Validation: S.X., A.A., O.C.S.; Visualisation: S.X., N.S.T., S.W.; Writing – original draft: S.X., A.K.D., S.W.; Writing – review & editing: All authors. All authors had full access to all the data in the study and accept responsibility for the decision to submit for publication. S.X. and S.W. verified the underlying data of the study.

## SUPPLEMENTARY INFORMATION

**Supplementary Data 1**. Supplemental methods, supplemental figures, and supplemental references.

**Supplementary Data 2**. Background sequence accessions and metadata

**Supplementary Data 3**. Sample metadata and sequencing metrics

**Supplementary Data 4**. Cholera cases reported by country and year

**Supplementary Data 5**. Median depth distribution by sample type from sub-sampling analysis

**Supplementary Data 6**. Median depth distribution by nucleotide position (4000-nt aggregated) after downsampling

**Supplementary Data 7**. Raw data file of maximum likelihood tree (n=1488)

**Supplementary Data 8**. Raw data file for posterior distribution of tMRCA of the AFR13 sub-lineage

**Supplementary Data 9**. Raw data file for root-to-tip divergence of all genomes (n=1499) and wave 3 genomes (n=961)

**Supplementary Data 10**. Raw data file BEAST MCC phylogeny of wave 3 genomes (n=961)

