## Supplementary Data 1 for "Whole genome sequencing and transmission analysis of *Vibrio cholerae* isolates from Eastern and Southern Africa: a genomic epidemiology study"

### Supplemental Methods

Bacterial culture and cholera confirmation

Specimens were streaked directly onto Thiosulfate Citrate Bile Salt sucrose (TCBS) agar and incubated overnight (18-24 hours) at 37°C. Immediately after inoculating the first TCBS plate, a pre-labeled APW vial was inoculated with the specimen and incubated for 4-6 hours at room temperature. After incubation, a second enriched specimen was inoculated on a TCBS plate and incubated overnight (18-24 hours) at 37°C. After overnight incubation, any cholera-like colonies were either tested via classical biochemical testing and polyvalent antisera serotyping [1] or selected with a sterile loop, resuspended in one to two drops of phosphate-buffered saline (PBS), and tested via dipstick or via polyvalent sera agglutination. All agglutination positive and dipstick-positive isolates, as well as any isolates considered cholera suspect (demonstrating the morphology of a cholera colony), were spotted (~50uL) on filter paper and/or inoculated in AFR1N1 agar (1% tryptone and 1% NaCl) for preservation.

DNA extraction and quantification

Each dried filter paper specimen was excised using sterile scissors and placed into a pre-labeled tube. 1 mL sterile 1X PBS was added to each sample tube and incubated for 10 minutes at room temperature. An additional 1 mL of sterile 1X PBS was then added to each sample and samples were immediately centrifuged (14,000 x g for 2 minutes) and the supernatant discarded. Subsequently, 150 µL of a 2% Chelex-100 solution (Bio-Rad) followed by 50 µL of sterile water was added to each sample. The samples were placed in a heating block at 100°C for 8 minutes and then centrifuged (14,000 x g for 2 minutes). The supernatant was transferred to a new microcentrifuge tube and either stored at -20°C or used in a PCR amplification reaction [2,3].

Glycerol stock preserved bacterial isolates were revived by taking a loop from the received glycerol stock and inoculating 3 mL Luria broth and incubating overnight (18-24 hours) at 37°C. The culture was then streaked onto a TCBS plate using a four-quadrant method and incubated overnight at 37°C. A single colony was selected and inoculated into 3 mL Luria broth and incubated shaking for 8 hours at 37°C. 1 mL of the turbid culture was used as input to QIAamp DNA Mini Kit (Qiagen) and centrifuged for 5 minutes at 5000 x g (7500 rpm). Volume of the pellet was calculated and Buffer ATL added to a total volume of 180 µL. The remaining steps of extraction were performed according to package directions for DNA purification from tissues.

We used conventional PCR to confirm toxigenic *V. cholerae* [4] and serogroup O1 [5] before quantification on the Qubit Fluorometer (Thermo Fisher) using the dsDNA High Sensitivity Kit.

Illumina library construction and sequencing

All samples were normalized to 0.6 ng/µL and Illumina sequencing libraries were prepared according to the Nextera DNA Flex Library Prep kit (Illumina). Library concentrations were measured using the Qubit High Sensitivity DNA Kit (Thermo Fisher), then normalized to a final concentration of 1ng/µL. Sample quality was then assessed on Caliper LabChip and/or Agilent BioAnalyzer, and samples were excluded if they were poor quality or had a concentration below 1ng/µL. All remaining samples were then combined into a single pool. Samples were sequenced on the Illumina NovaSeq platform with 2x150 bp paired-end reads. The entire pool was run in duplicate on an Illumina NovaSeq.

Oxford Nanopore library construction and sequencing

Oxford Nanopore library preparation and sequencing was performed using a starting input amount of 350-1300 ng in 48 µL volume. Libraries were prepared following the SQK-LSK109 library preparation kit from Oxford Nanopore Technologies, with minor modifications as described in Ekeng et al. [6] The final pooled library was eluted in 15 µL Elution Buffer and 190 ng was diluted to 12 µL for loading onto the MinION flow cell. The MinION was run for a total of 48 hours per run and the resulting data was basecalled using Guppy version 3.0.3 with model dna_r9.4.1_450bps_fast.cfg. Adapter removal and demultiplexing was performed with Porechop version 0.3.2 [7] and reads were filtered using Filtlong version 0.2.1 [8] with the following options: '--keep_percent 90 --target_bases 800000000.'

Reference-based genome assembly

Illumina paired-end reads were aligned against *V. cholerae* O1 El Tor N16961 (accession: AE003852/AE003853). For each sample, paired-end reads from two lanes were mapped against the reference genome using BWA version 0.7.17 [9] to produce a SAM file. The SAM files from two lanes were converted to BAM files, sorted and merged using samtools version 1.13-17 [10]. Picard version 2.26.0 [11] was used to mark duplicates in the merged BAM file and variants were called using bcftools version 1.13.35 [12]. Variants were then filtered to obtain VCF files with minimum variant quality score of 20 and a minimum mapping score of 30. Variants were also required to be present in at least 75% of reads mapped and present on both strands (≥2 read depth per stand) [13]. The criteria for inclusion of the genome for subsequent analysis were: (1) median coverage number across all positions on the genome greater than 20; and (2) at least 97% coverage of the reference genome (<%3 ambiguous bases). The complete Illumina genome assembly pipeline used in this study is publicly available at https://github.com/HopkinsIDD/illumina-vc.

For Oxford Nanopore data, reference-based genome assembly was performed as described in Ekeng et al. [6]. The complete Oxford Nanopore genome assembly pipeline used in this study is publicly available at https://github.com/HopkinsIDD/minion-vc.

Visualization of coverage depth across genomes

We compared the reference genome coverage across sample types for all Illumina-sequenced samples. First, we used seqtk version 1.2-r94 [14] to downsample the raw sequencing reads to 1 million reads per sample. We then used samtools version 1.13-17 [10] to calculate the read depth at each position across the genome. To determine the distribution of overall coverage depth by sample type, we calculated the median coverage depth across the genome for each sample. To visualize the distribution of coverage depth for each position in the genome by sample type, we calculated the median, 20th and 80th percentile coverage depth at each nucleotide position across all samples within each sample type group [15]. We then plotted the mean of each of these metrics within a 4000-nt sliding window.

Assembling the background dataset

To generate the background dataset listed in **appendix 2**, we downloaded publicly available whole genome *V. cholerae* sequences as of July 2021. We selected these genomes to capture as much global diversity as possible, with an emphasis on capturing published genomes from sub-Saharan Africa. In most cases, sequences were published as reference-based assemblies generated using the same N16961 reference as used in this study [6,13,16]. However, in some cases we did not have access to assemblies and had to assemble genomes from raw Illumina FASTQ files or contigs. Sequences published as raw Illumina paired-end reads (accession: PRJEB30604) were downloaded and run through the assembly pipeline described above. Sequences published as contigs (accession: PRJNA729102) were processed using snippy version 4.6.0 [17]. The snippy software was used to produce a BAM file aligned to the N16961 reference, which was then subjected to the variant calling process described above.

Maximum likelihood estimation

We concatenated the 114 genomes generated in this study with the 1,385 genomes in our background dataset, described above, resulting in a total dataset of 1,499 genomes—all of which have been assembled using the same N16961 reference sequence. We used the same method for masking recombinant sites on sequences generated on the Illumina and Oxford Nanopore platforms, as well as on previously published genomes. Masking was a two-part process as described in Weill et al. [16]: (1) known recombinant regions were masked using a custom GFF file (see https://figshare.com/s/d6c1c6f02eac0c9c871e); (2) genomes were concatenated into a pseudo-alignment and additional sites were masked using gubbins version 2.3.4 [18]. A maximum likelihood tree was then constructed on the masked SNP alignment from these 1,499 genomes using IQ-TREE version 1.6.12 [19,20] with a General Time Reversible (GTR) substitution model [16] and 1000 bootstrap iterations [6]. We later reran the maximum likelihood estimation on the outlier-removed dataset of 1,488 genomes (see below). Figtree version 1.4.4 [21] and R package ggtree version 3.4.0 [22] and treeio version 1.20.2 [23] were used for tree visualization.

Root-to-tip regression for identifying outliers

The maximum likelihood trees were examined for the degree of temporal signal using TempEst v1.5.3 [24]. We identified 11 (all previously published) outlier sequences whose genetic divergence and sampling date were incongruent under a linear regression of root-to-tip divergence (residual >0.01) and removed these from our dataset (**Supplementary Figure 1**). For samples where we did not have a specific date (collection year only), we used the middle of the collection year for this analysis. Maximum likelihood estimation was rerun as described above, resulting in a final tree with 1,488 sequences and a wave 3-specific tree (see below) with 961 sequences (**appendix 2**).

BEAST analysis for wave 3 samples

Evolutionary and temporal dynamics of wave 3 sequences were reconstructed with a Bayesian phylogenetic approach using Markov chain Monte Carlo (MCMC) available via the BEAST version 1.10.5 package [25] and the high-performance computational capabilities of the Biowulf Linux cluster at the National Institutes of Health, Bethesda, MD, USA [26]. A general time-reversible substitution model with gamma-distributed rate heterogeneity (GTR + gamma) was used under a strict molecular clock with a Skyline coalescent prior [16,27]. For sequences with only collection year available, the lack of tip date precision was accommodated by sampling uniformly across a one-year window from 1st January to 30th December.

Ten independent Markov chain Monte Carlo chains with three checkpoints [28] were run for 200 million steps and sampled every 20,000th generation, with at least 10% of the generations discarded as chain burn-in. All analyses were performed using the BEAGLE library to enhance computation speed [29,30]. Convergence and mixing of the chains were inspected using Tracer version 1.7.3 [31]; all continuous parameters yielded effective sample sizes greater than 200. A maximum clade credibility tree was summarized using TreeAnnotator version 1.10.5 [25] and visualization of the tree with annotations was performed with FigTree version 1.4.4 [21] (**Supplementary Figure 2**). Distribution of the time to the most recent common ancestor (tMRCA) for lineage AFR13 was performed using TreeStat version 1.10.5 [32].

Geospatial data

Country and sub-country shapefiles used in **Figure 4** were obtained in R from *gadm* version 4.1 [33]. The water shapefile was from World Wild Life global lakes and wetlands database [34] (retrieved March 3, 2023). R packages *sf* version 1.0-9, *ggspatial* version 1.1.6, *ggthemes* version 4.2.4, *geodata* version 0.4-11, *raster* version 3.6-3, and *geos* version 0.2.2 were used for making maps [35–40]. In all geospatial analysis, we mapped samples to the centroid of their collection country if no sub-country location information was provided.

### Supplemental Figure 1

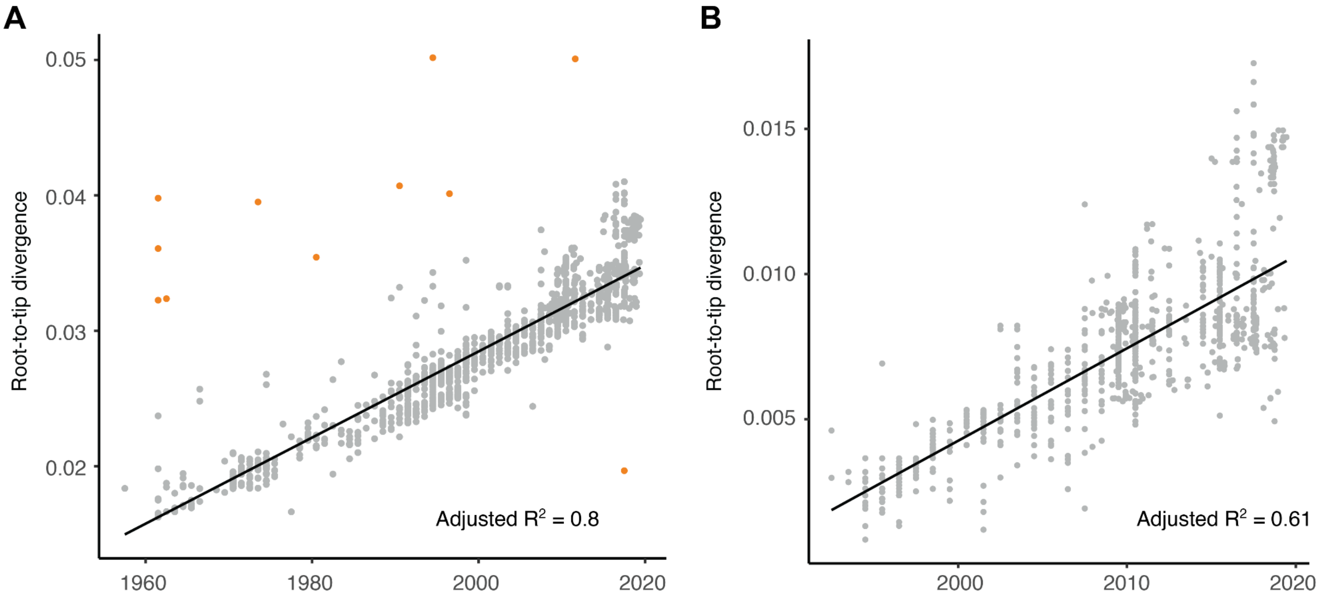

**Molecular clock validation.** (**A**) Root-to-tip regression of background genomes plus all 114 genomes generated in this study (n = 1499). Orange dots: outliers identified in previously published genomes. Samples represented by gray dots were included in the maximum likelihood tree analysis (N = 1488). (**B**) Root-to-tip regression of 961 wave 3 genomes included in the BEAST analysis (see **appendix 9**).

### Supplemental Figure 2

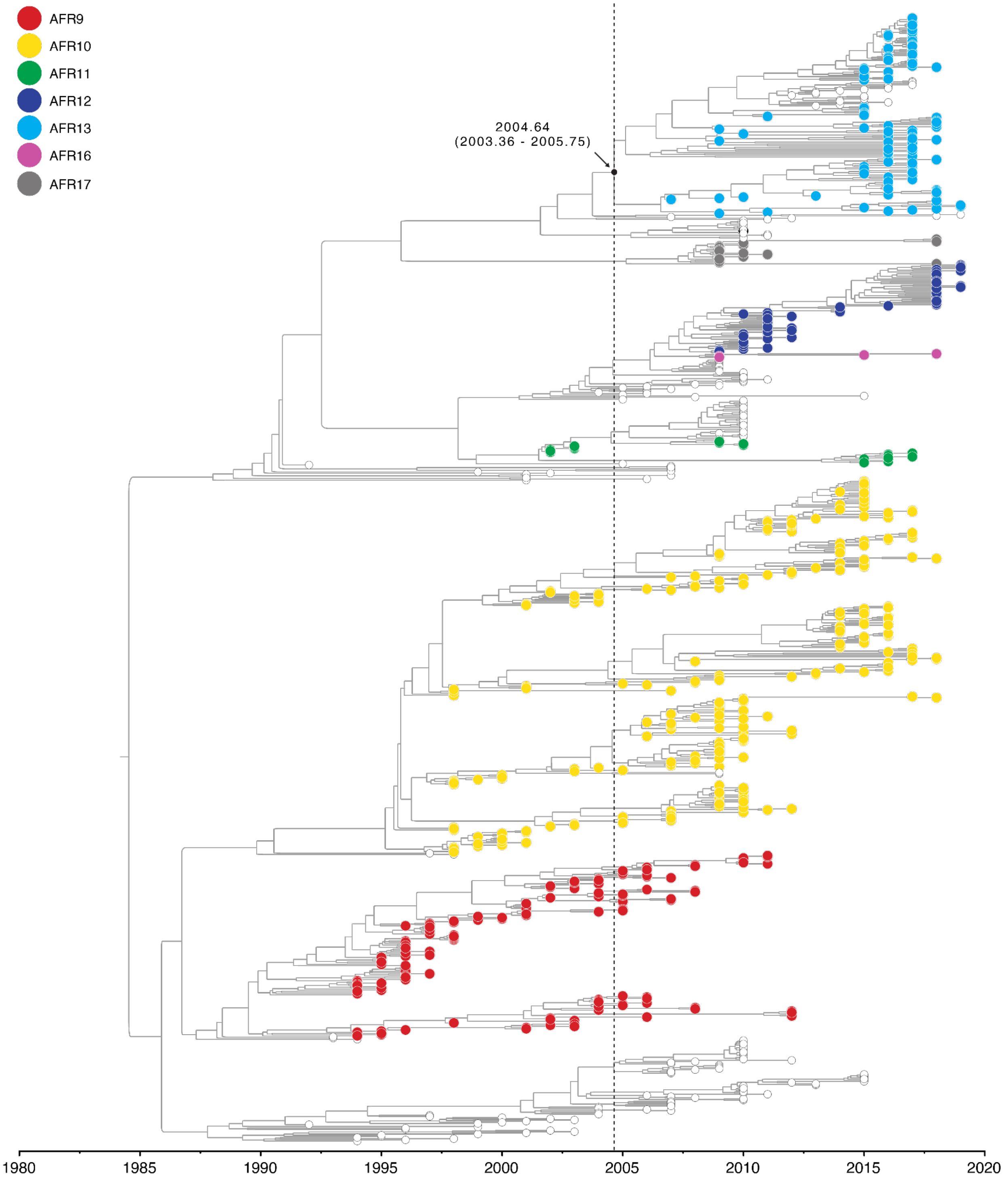

**Wave 3 maximum clade credibility tree.** Maximum clade credibility tree depicting time-scaled phylogeny of *V. cholerae* wave 3 (see **appendix 10**). Tips are colored by lineage, with non-AFR lineages shown as smaller white tips. Median tMRCA of the AFR13 lineage is indicated on tree (see also **Figure 3C**).
